# Polygenic Risk for Substance-Related Traits Predicts Substance Use Onset and Progression: Sex and Population Group Differences

**DOI:** 10.1101/2022.09.29.22280477

**Authors:** Henry R. Kranzler, Richard Feinn, Heng Xu, Brendan L. Ho, Divya Saini, Olivia R. Nicastro, Anya Jacoby, Sylvanus Toikumo, Joel Gelernter, Emily E. Hartwell, Rachel L. Kember

**Affiliations:** Department of Psychiatry, University of Pennsylvania Perelman School of Medicine, Philadelphia, PA 19104; Mental Illness Research, Education and Clinical Center, Veterans Integrated Service Network 4, Crescenz Veterans Affairs Medical Center, Philadelphia, PA 19104; Department of Medical Sciences, Frank H. Netter School of Medicine at Quinnipiac University, North Haven, CT 06473; Department of Psychiatry, Yale University School of Medicine, New Haven, CT 06511 and VA Connecticut Healthcare System, West Haven, CT 06516

**Author notes:** Correspondence to Henry R. Kranzler, M.D., Center for Studies of Addiction, 3535 Market Street, Suite 500, Philadelphia, PA 19104.

## Abstract

**Background:** Charting the clinical course of substance use disorders (SUDs) to identify etiologic contributors to milestone onset and progression could inform intervention efforts.

**Methods:** We calculated polygenic risk scores (PRS) in 5,692 European-ancestry individuals (EUR) (56.2% male) and 4,918 African-ancestry (AFR) individuals (54.9% male) using genome-wide association studies (GWAS) of alcohol use disorder (AUD), opioid use disorder (OUD), and smoking trajectory (SMK). Using Cox regression, we examined the association of polygenic risk with age of first substance use, regular use, reported problems, and dependence diagnosis and with progression from regular use to onset of problems and dependence.

**Results:** EUR and males reported earlier onset and shorter progression times than AFR and females, respectively. Among EUR, higher AUD PRS predicted earlier onset and more rapid progression to alcohol-related milestones (*p*’s<0.0001) and although a stronger moderator of problem onset among females (*p*=0.0165), it was more predictive of the progression to problems among males (*p*=0.0054). OUD and SMK PRS in EUR also predicted earlier onset of the respective milestones (*p*’s=0.0002). Among AFR, where power is lower, AUD PRS predicted age of regular alcohol use (*p*=0.039) and dependence (*p*=0.001) and progression from regular use to diagnosis (*p*=0.045), while SMK PRS predicted earlier age of initiation (*p*=0.036).

**Conclusions:** Genetic risk for SUDs predicts milestones and symptom progression in EUR and, to a lesser extent, among AFR. Larger, diverse discovery GWAS and target samples are needed to enhance the power of PRS to personalize interventions for individuals at genetic risk of serious substance-related outcomes.

**Disclosure:** Dr. Kranzler is a member of advisory boards for Dicerna Pharmaceuticals, Sophrosyne Pharmaceuticals, and Enthion Pharmaceuticals; a consultant to Sobrera Pharmaceuticals; the recipient of research funding and medication supplies for an investigator-initiated study from Alkermes; and a member of the American Society of Clinical Psychopharmacology’s Alcohol Clinical Trials Initiative, which was supported in the last three years by Alkermes, Dicerna, Ethypharm, Lundbeck, Mitsubishi, and Otsuka. Drs. Gelernter and Kranzler hold U.S. Patent 10,900,082: Genotype-guided Dosing of Opioid Receptor Agonists, 26 Jan. 2021. The other authors have no disclosures to make.

## Introduction

Substance use disorders (SUDs), characterized by the chronic use of alcohol or drugs, are common among adolescents and adults and result in clinically significant social impairments and medical and psychiatric disorders.^1^ SUDs develop in stages following initial substance use, often progressing across a series of sequential transitions, which can be conceptualized as a continuous trajectory marked by milestones of escalating use or severity.^2^ Charting the clinical course of SUDs using these developmental events can help to elucidate the factors that underlie symptom progression, a key example of which is the transition from regular substance use to substance dependence. The timing of milestones could also provide a personalized assessment of an individual’s risk of symptom progression^3^ and a more precise time window for a targeted intervention aimed at preventing the progression to a subsequent milestone.^4^

The age at which substance-related milestones occur and the rate of progression through them are influenced by both genetic and environmental factors.^4^ These risk factors may be reflected in population-group differences in the initiation of substance use and the progression across substance-related milestones.^5^ For example, Black individuals report first consuming alcohol and tobacco products and initiating regular drinking and binge drinking, and use of illicit drugs, later than White individuals.^6,7,8^

Although Black adolescents had a significantly lower risk of transitioning to regular use of alcohol and illicit drugs,^8^ Black adults 30 years of age and older have also been shown to have a more rapid progression from regular alcohol use to regular drinking and intoxication than White adults.^9^ These differences occur in the context of a general paucity of studies of population-group differences in substance-related symptom progression.^10^ There is a similar dearth of findings on population group differences in age-of-onset and progression measures for opioids, though in one study Black individuals had a more rapid progression to opioid dependence (OD) than White individuals.^11^

Sex may also influence the developmental course of substance-related traits. In monozygotic and dizygotic twins, the initiation of alcohol and tobacco use occurred earlier among males than females with earlier initiation of use associated with a significantly increased likelihood of developing dependence on these substances^4^. In contrast, in another twin study, between-twin comparisons of the rate of progression through alcohol-related milestones showed no overall pattern of sex differences.^2^ Thus, the relationship of sex to alcohol-related milestones and progression in SUDs is not fully understood. A controversial question in relation to sex differences is the validity of the phenomenon of telescoping, which posits that despite their later initiation of substance use, women manifest substance-related problems sooner than men ^.2^

Risk for SUDs is highly polygenic, involving potentially thousands of individual variants, each accounting for a very small proportion of trait variance. Polygenic risk scores (PRS) sum data from multiple genetic variants and account for greater proportions of trait variance than single polymorphisms.^13^ PRS are useful in evaluating the risk for disease progression in diverse medical disorders, including breast cancer^14^ and rheumatoid arthritis,^15^ and the prediction of sudden death in individuals with coronary disease.^16^ A recent large study of EUR in the UK Biobank showed the utility of a prostate cancer genetic risk score for triaging patients in primary care. Men in the highest quintile of risk had a prostate cancer incidence of 8.1% and could be fast-tracked for further investigation, while the incidence among those in the lowest risk quintile was <1% and they could more safely avoid invasive investigation.^17^ Thus, there currently are clinical applications of PRS to differentiate individuals based on their genetic risk for a disease.

For SUDs, a PRS for alcohol dependence (AD) was associated with the progression from onset of regular drinking to AD in a European-ancestry (EUR) sample.^18^ The ability to quantify the genetic risk of symptom progression could help to identify individuals at highest risk of developing more severe milestones (e.g., alcohol-related problems or AD) and who could benefit most from intensive interventions.

Here, we examined the previously reported association of an AD PRS with the progression from onset of regular drinking to onset of AD in EUR.^18^ We then extend the analyses to include additional milestones and measures of progression of alcohol-related symptoms and conduct parallel analyses of opioid-related and smoking-related traits. Finally, we refine our understanding of the effects of sex and population group differences on these features by studying males and females and EUR and African-ancestry (AFR) participants, all of whom are well represented in our sample, which was recruited and deeply phenotyped for genetic studies of SUDs.^19^

## Methods

### Discovery Samples

We used GWAS summary statistics for alcohol use disorder (AUD),^20^ opioid use disorder (OUD),^21^ and smoking trajectories (SMK)^22^ as discovery samples for calculating PRS in the Yale-Penn sample. The three discovery samples are described in detail in Supplemental Methods.

To compare the power among the discovery samples, we calculated the genomic inflation factor (λgc). In the absence of population structure, λgc is a function of sample size and the number of causal variants and thus reflects power to detect significant SNP-trait associations.^23^ We calculated λgc using the expected median of SNP test statistics from each set of GWAS summary statistics, with greater λgc denoting higher predictive power. These showed that for all three traits EUR samples had better predictive power than AFR samples. For AUD, the λgc was 1.100 for EUR and 1.034 for AFR, while for OUD the respective values were 1.112 and 1.028, and for SMK they were 1.336 and 1.094.

### Target Sample

We calculated PRS in the Yale-Penn sample, which comprises 16,715 individuals recruited at five U.S. sites for genetic studies of dependence on cocaine, opioids, and alcohol.^19^

Of this number, genome-wide genotype data are available for 10,610 individuals, including 5,692 EUR (56.2% male) and 4,918 AFR (54.9% male) (Table 1). The study was approved by the institutional review board at each site and participants gave written informed consent for data collection. Cases were identified through addiction treatment facilities, inpatient and outpatient psychiatric services, and posters and advertisements in local media. Although some cocaine- or opioid-dependent individuals were recruited as probands of small nuclear families, data from family members are excluded in analyses presented here. Unaffected controls were recruited from non-psychiatric medical settings and through advertisements.

**Table 1.**
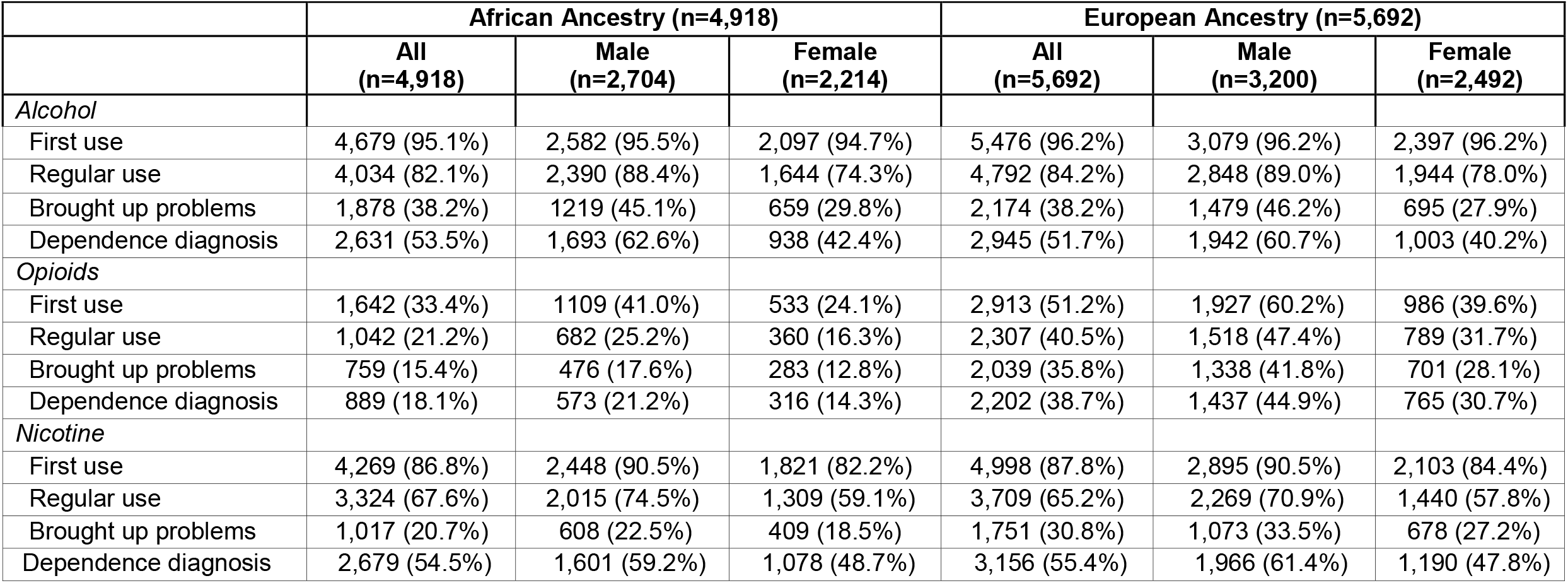
Prevalence of Substance Use Disorder Milestones by Ancestral Group and Sex

### Semi-Structured Assessment for Drug Dependence and Alcoholism (SSADDA)

The SSADDA is a comprehensive psychiatric interview that comprises 24 modules assessing the physical, psychological, social, and psychiatric manifestations of SUDs, psychiatric disorders, and environmental covariates considered likely to have an impact on SUDs. The SSADDA’s semi-structured format, accompanied by the rigorous training and quality control procedures used in the Yale-Penn sample,^24^ allow a carefully trained non-clinician interviewer to assess diagnostic criteria and disorders and their ages of onset, to yield DSM-IV diagnoses of AD, OD, and nicotine dependence (ND). Ascertainment of ages of onset of the different milestones is done using questions that elicit estimated ages (e.g., of initiation of substance and regular substance use) and clustering of criteria within a 12-month period (for DSM-IV diagnoses).

### Genotyping, Imputation, and Polygenic Risk Scores

Yale-Penn samples were genotyped in three batches using the Illumina HumanOmni1-Quad microarray, the Illumina HumanCoreExome array, or the Illumina Multi-Ethnic Global array. Genotype data were filtered for individual call rates and excessive heterozygosity using PLINK v1.9 and were imputed using the Michigan Imputation Server^25^ and the Haplotype Reference Consortium Panel.^26^

PRS were calculated for AUD,^20^ OUD,^21^ and SMK^22^ using Polygenic Risk Scores– Continuous Shrinkage software (PRS-CS)^27^ and the 1000 Genomes Project phase 3 AFR and EUR samples for estimates of linkage disequilibrium. Global shrinkage parameters were obtained from each set of summary statistics by the PRS-CS package and effective sample sizes were used to calculate the final PRS. We used matched, genetically determined, ancestral summary statistics (e.g., an AFR GWAS for AUD was used to calculate AUD PRS in AFR Yale-Penn individuals). Whereas PRS calculated with the effective or actual sample sizes did not differ substantially, we report findings using effective sample sizes.

### Statistical Analysis

We used PROC PHREG in SAS v9.4 to run Cox proportional hazards models for each of three substances (alcohol, opioids, and smoking) and the following four measures: age of first use, age of regular use, age of first bringing up problems with a healthcare professional, and age of diagnosis of the disorder (AD, OD, and ND). Using these milestones, we also examined two measures of progression: time from age of regular use to age of first bringing up problems and time from age of regular use to age of diagnosis. Analyses were conducted separately for EUR and AFR, for which there are comparable numbers in the Yale-Penn sample. Similarly, analyses were conducted separately for females and males within each ancestry group, also roughly equally distributed in the target sample.

All models included the respective PRS, age, and the first 10 principal ancestral components as covariates. In analyses that did not examine sex as a factor, sex was included as a covariate. The models for the progression outcomes also included the age of regular use as a covariate. Because the proportional hazards assumption for this covariate was violated in the progression models for all three substances, we added an interaction term for age of regular use for the two progression measures, which was significant in all progression models. Thus, we assessed the impact of age of onset of regular use on the two progression outcomes by decomposing the effects for participants with early onset (≤18 years) and late onset (>18 years) of regular use.^18^

For individuals who reported never having experienced a specific event, data were censored for the event in the Cox models and the age at interview was substituted for the missing age of onset. Thus, the age-of-onset outcomes for all substances have the same sample size. For the two progression outcomes, only individuals who reported regular use were included in the analysis, as censoring that age of onset would distort the analysis. Thus, the sample sizes for the two progression outcomes vary by substance.

For effect sizes, we report the hazard ratio (HR) with 95% confidence intervals, reflecting the change in hazard for a one-standard-deviation increase in PRS. For individual milestones, a HR>1.0 reflects a greater likelihood that the event will occur as PRS increases. Similarly, with progression (or latency) measures, there is a greater likelihood of progression from onset of regular substance use to either first reported problems or first dependence diagnosis as PRS increases. We also report p-values and adjusted p-values using the Hommel correction for multiple testing, with adjustments made separately by population group and substance. We consider PRS as a significant predictor when the p-value, adjusted for multiple comparisons, is <0.05.

## Results

### Rates of Endorsement of Substance Use Milestones

Table 1 shows the prevalence of the milestones of first use, regular use, first reporting of problems, and onset of dependence, for alcohol, opioids, and nicotine in the Yale-Penn sample. More than 95% of both population groups reported ever having used alcohol and over 80% reported ever having used alcohol regularly. Lifetime opioid use was less common among AFR (33%) than EUR (51%), as was regular opioid use (21% vs. 41%). In both groups, nearly 90% of individuals reported having smoked more than 100 cigarettes lifetime, with over 65% endorsing regular smoking. In both AFR and EUR, the event endorsed least commonly for all three substances was having brought up problems to a healthcare professional, which ranged from 15%-31% in AFR and 31%-38% in EUR.

Most individuals in both population groups met DSM-IV criteria for AD and ND, while the prevalence of OD was 39% among EUR and less than half that among AFR (18%). The high rate of SUD diagnoses in the Yale-Penn sample reflects its ascertainment for studies of SUD genetics.^19^ The sample prevalence of all three SUD diagnoses is lower among women and AFR and the onset of all milestones earlier than among men and EUR, respectively. Similarly, EUR and males reported earlier onset and shorter progression times for all substances than AFR and women, respectively (Table 2).

**Table 2.**
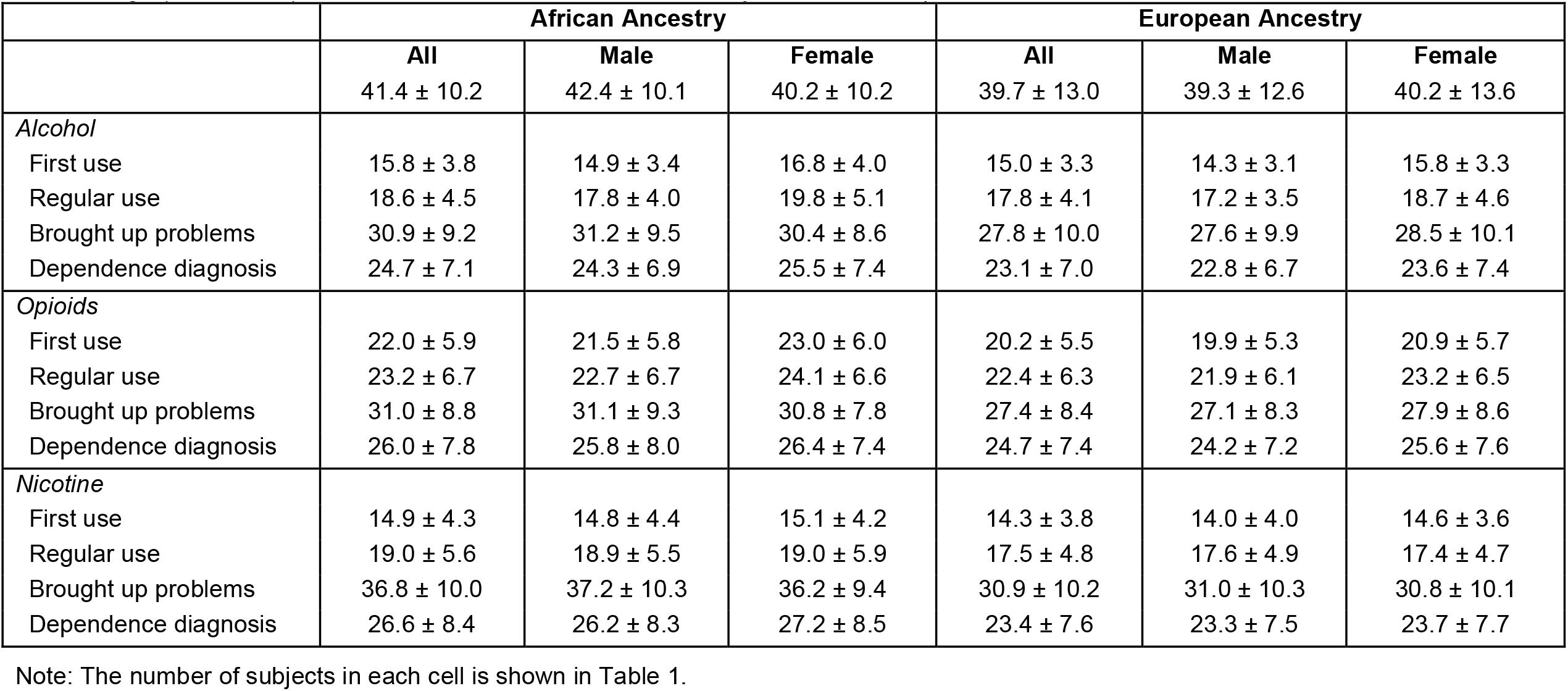
Age (Mean ± SD) of Substance Use Disorder Milestones by Ancestral Group and Sex

|  | **African Ancestry** |  |  | **European Ancestry** |  |  |
| --- | --- | --- | --- | --- | --- | --- |
|  | **All** | **Male** | **Female** | **All** | **Male** | **Female** |
| | 41.4 ± 10.2 | 42.4 ± 10.1 | 40.2 ± 10.2 | 39.7 ± 13.0 | 39.3 ± 12.6 | 40.2 ± 13.6 |
| *Alcohol* |  |  |  |  |  |  |
| First use | 15.8 ± 3.8 | 14.9 ± 3.4 | 16.8 ± 4.0 | 15.0 ± 3.3 | 14.3 ± 3.1 | 15.8 ± 3.3 |
| Regular use | 18.6 ± 4.5 | 17.8 ± 4.0 | 19.8 ± 5.1 | 17.8 ± 4.1 | 17.2 ± 3.5 | 18.7 ± 4.6 |
| Brought up problems | 30.9 ± 9.2 | 31.2 ± 9.5 | 30.4 ± 8.6 | 27.8 ± 10.0 | 27.6 ± 9.9 | 28.5 ± 10.1 |
| Dependence diagnosis | 24.7 ± 7.1 | 24.3 ± 6.9 | 25.5 ± 7.4 | 23.1 ± 7.0 | 22.8 ± 6.7 | 23.6 ± 7.4 |
| *Opioids* |  |  |  |  |  |  |
| First use | 22.0 ± 5.9 | 21.5 ± 5.8 | 23.0 ± 6.0 | 20.2 ± 5.5 | 19.9 ± 5.3 | 20.9 ± 5.7 |
| Regular use | 23.2 ± 6.7 | 22.7 ± 6.7 | 24.1 ± 6.6 | 22.4 ± 6.3 | 21.9 ± 6.1 | 23.2 ± 6.5 |
| Brought up problems | 31.0 ± 8.8 | 31.1 ± 9.3 | 30.8 ± 7.8 | 27.4 ± 8.4 | 27.1 ± 8.3 | 27.9 ± 8.6 |
| Dependence diagnosis | 26.0 ± 7.8 | 25.8 ± 8.0 | 26.4 ± 7.4 | 24.7 ± 7.4 | 24.2 ± 7.2 | 25.6 ± 7.6 |
| *Nicotine* |  |  |  |  |  |  |
| First use | 14.9 ± 4.3 | 14.8 ± 4.4 | 15.1 ± 4.2 | 14.3 ± 3.8 | 14.0 ± 4.0 | 14.6 ± 3.6 |
| Regular use | 19.0 ± 5.6 | 18.9 ± 5.5 | 19.0 ± 5.9 | 17.5 ± 4.8 | 17.6 ± 4.9 | 17.4 ± 4.7 |
| Brought up problems | 36.8 ± 10.0 | 37.2 ± 10.3 | 36.2 ± 9.4 | 30.9 ± 10.2 | 31.0 ± 10.3 | 30.8 ± 10.1 |
| Dependence diagnosis | 26.6 ± 8.4 | 26.2 ± 8.3 | 27.2 ± 8.5 | 23.4 ± 7.6 | 23.3 ± 7.5 | 23.7 ± 7.7 |

Note: The number of subjects in each cell is shown in Table 1.

### PRS Effects on Alcohol-related Milestones and Symptom Progression

Table 3 and Figure 1 (upper two panels) show the Cox regression model of the effects of the AUD PRS on alcohol-related milestones by population group. Significant associations reflect a younger age of onset and a shorter latency between milestones as a function of increasing PRS. Among EUR, AUD PRS was a significant predictor (adjusted p-value [*p*_*adj*_]<0.001) of all four milestones, with HRs ranging from 1.06 (age of first use and age of regular use) to 1.19 (age at which alcohol-related problems were brought up to a healthcare professional). The AUD PRS was also a significant predictor of the progression from age of regular alcohol use both to bringing up alcohol-related problems (HR=1.14, *p*_*adj*_<0.001) and age of AD diagnosis (HR=1.10, *p*_*adj*_<0.001) (Figure 2).

**Table 3.**
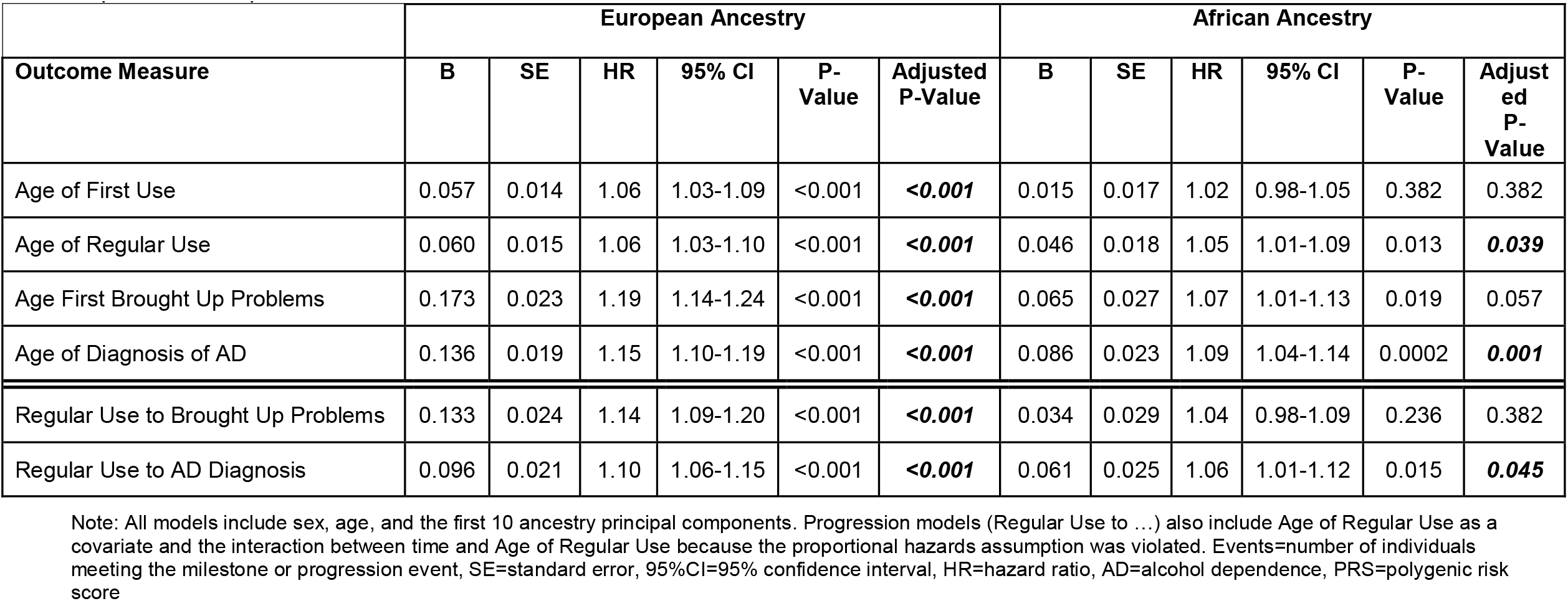
Effect of Alcohol Use Disorder Polygenic Risk Score on Age of Onset and Progression of Alcohol-Related Measures by Population Group

**Figure 1.**
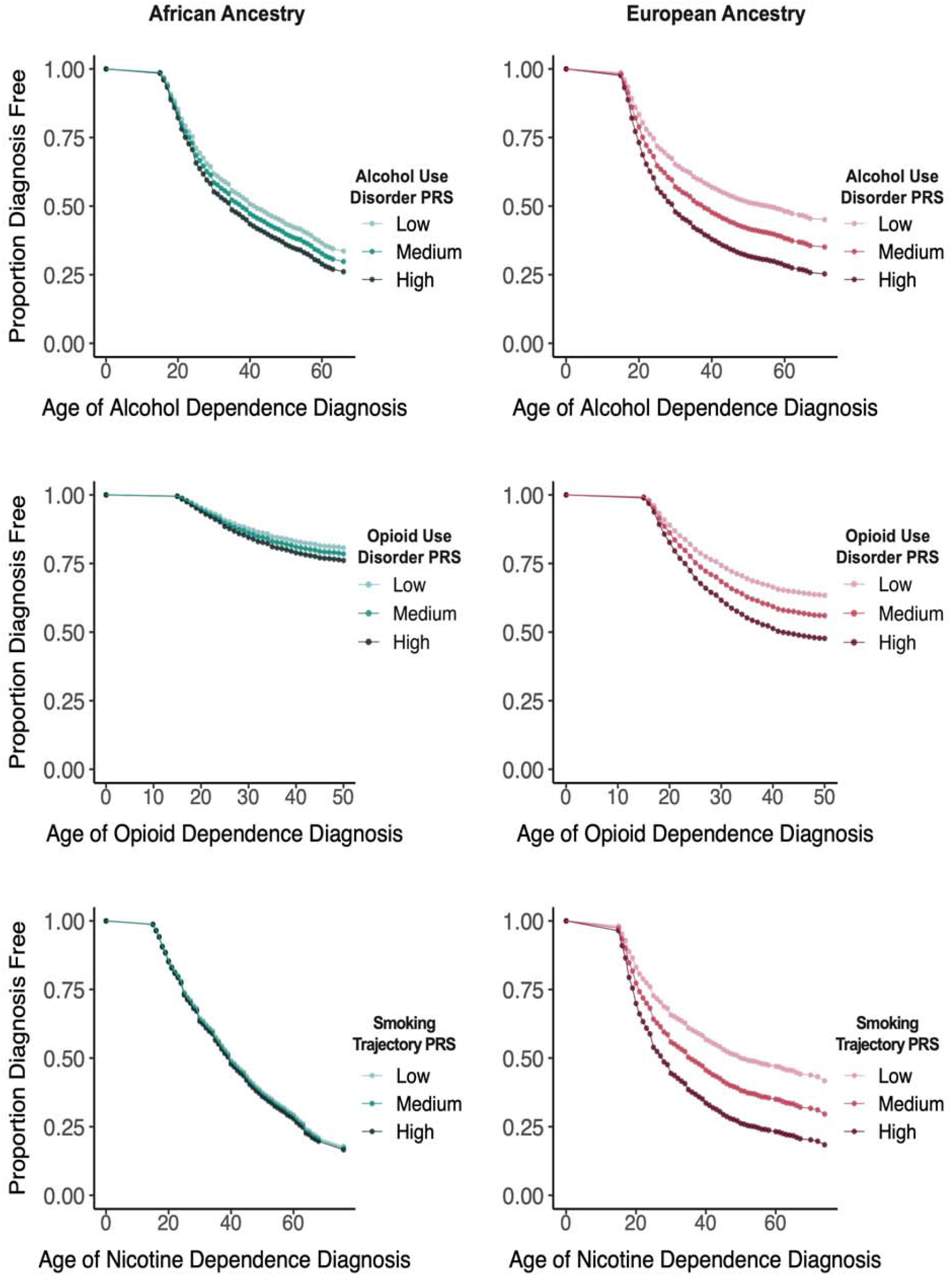
Age of Alcohol, Opioid, and Nicotine Dependence Diagnoses by European or African Ancestry and Low, Medium, and High Polygenic Risk Scores for Alcohol Use Disorder, Opioid Use Disorder, and Smoking Trajectory, Respectively

**Figure 2.**
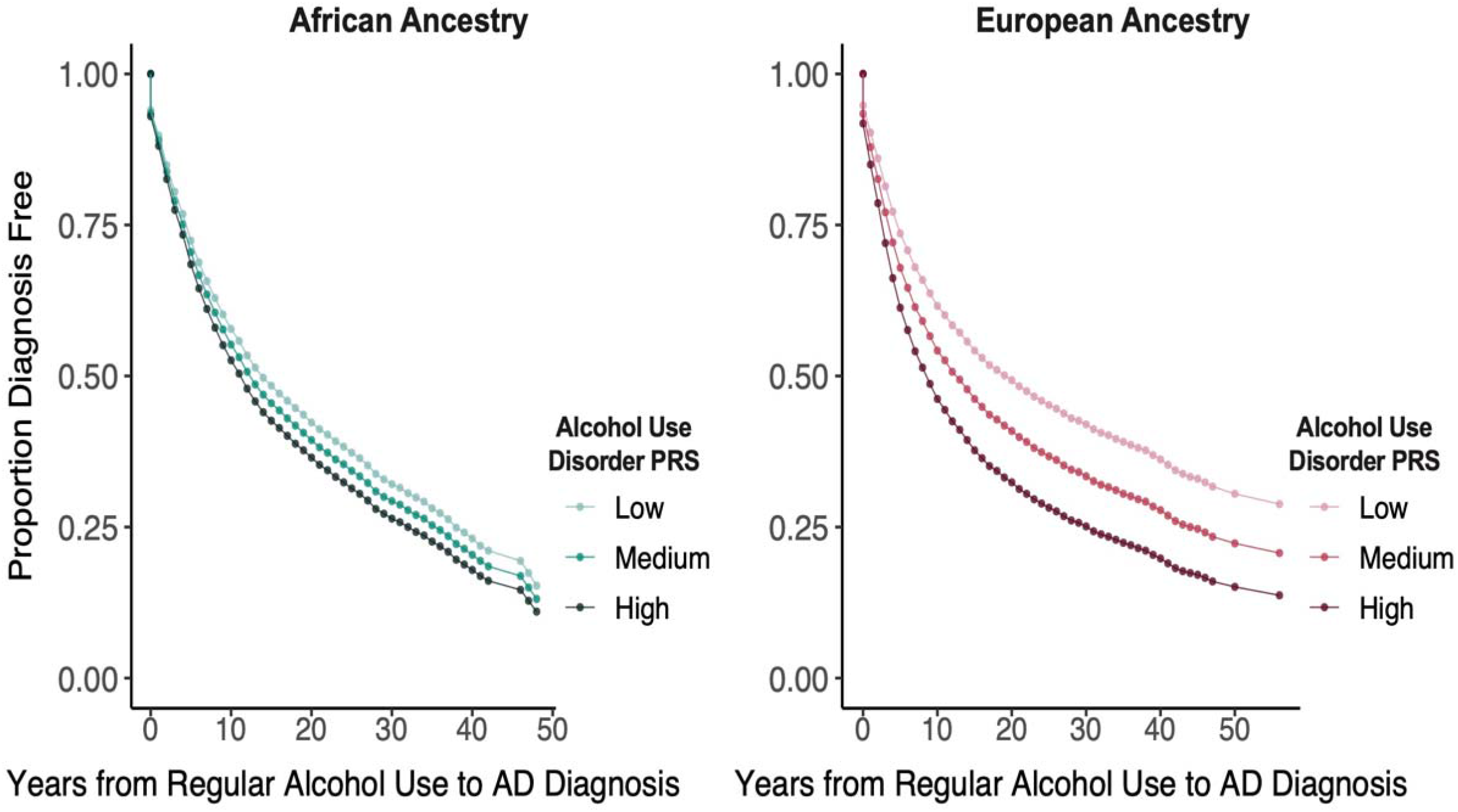
Years from Regular Alcohol Use to Alcohol Dependence Diagnosis by European or African Ancestry and Low, Medium, and High Alcohol Use Disorder Polygenic Risk Scores

Among AFR, the AUD PRS significantly predicted both age of regular alcohol use (HR=1.05, *p*_*adj*_=0.039) and age of AD diagnosis (HR=1.09, *p*_*adj*_=0.001) (Table 3). It was also a significant predictor of the progression from age of regular alcohol use to age of AD diagnosis (HR=1.06, *p*_*adj*_=0.045) (Figure 2).

In analyses stratified by age-of-onset of regular drinking (Supplemental Table 1), in both population groups, early onset of regular drinking (≤18 years) was associated with more rapid progression both to age of first reported alcohol-related problems and to age of initial AD diagnosis. Among EUR, the AUD PRS, irrespective of the age of onset of regular drinking, significantly predicted the progression to first reported alcohol-related problems and to an AD diagnosis (HRs = 1.09-1.18). Among AFR, the AUD PRS was positively associated with both progression measures (HRs=1.02-1.11), though the effect was significant only for progression to an AD diagnosis and only among individuals with a late onset of regular drinking (>18 years).

A sex-stratified analysis among EUR (Table 4 and Figure 3) showed that the association of AUD PRS with milestones was greater among women (HRs=1.06-1.30) than men (HRs=1.06-1.15). There was also a significant interaction effect of sex by AUD PRS on the age at which alcohol-related problems were raised with a health professional (*p*_*adj*_=0.0165). The genetic risk for AUD was a stronger moderator in females than males despite men having brought up problems earlier than women. This is evidenced by greater separation between survival curves for low, medium, and high PRS tertiles among women than men (Figure 3).

**Table 4.**
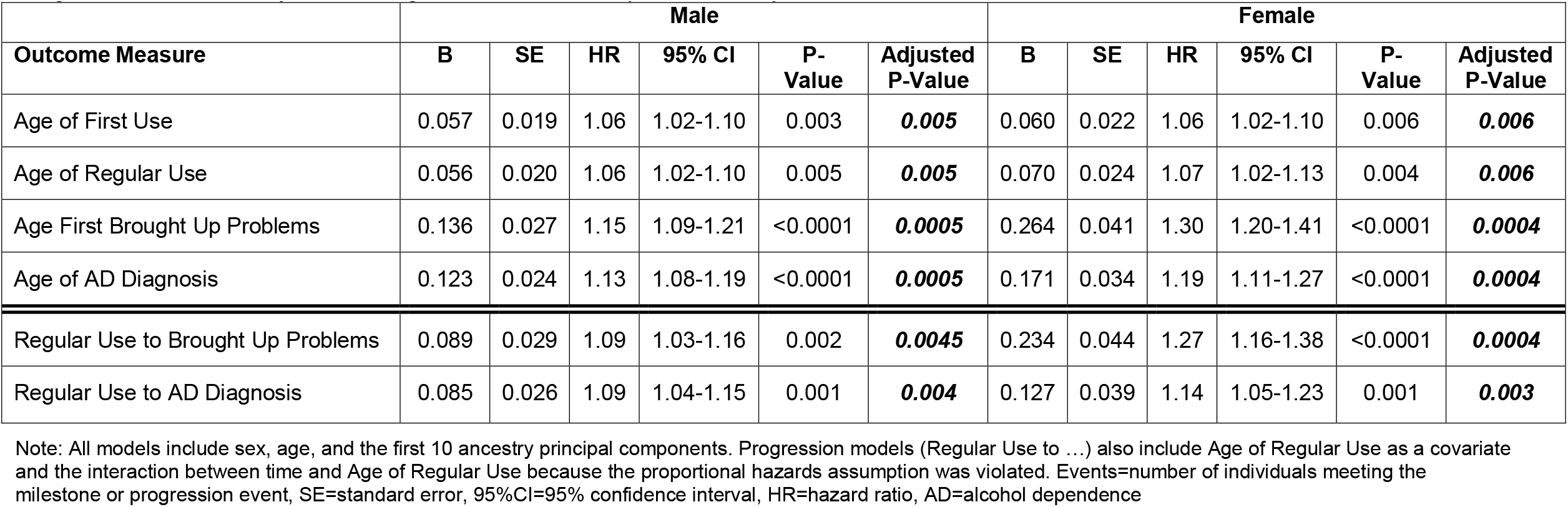
Effect of Alcohol Use Disorder Polygenic Risk Score in Cox Proportional Hazard Models of Alcohol-Related Time-to-Event and Progression Measures by Sex among Individuals of European Ancestry

**Figure 3.**
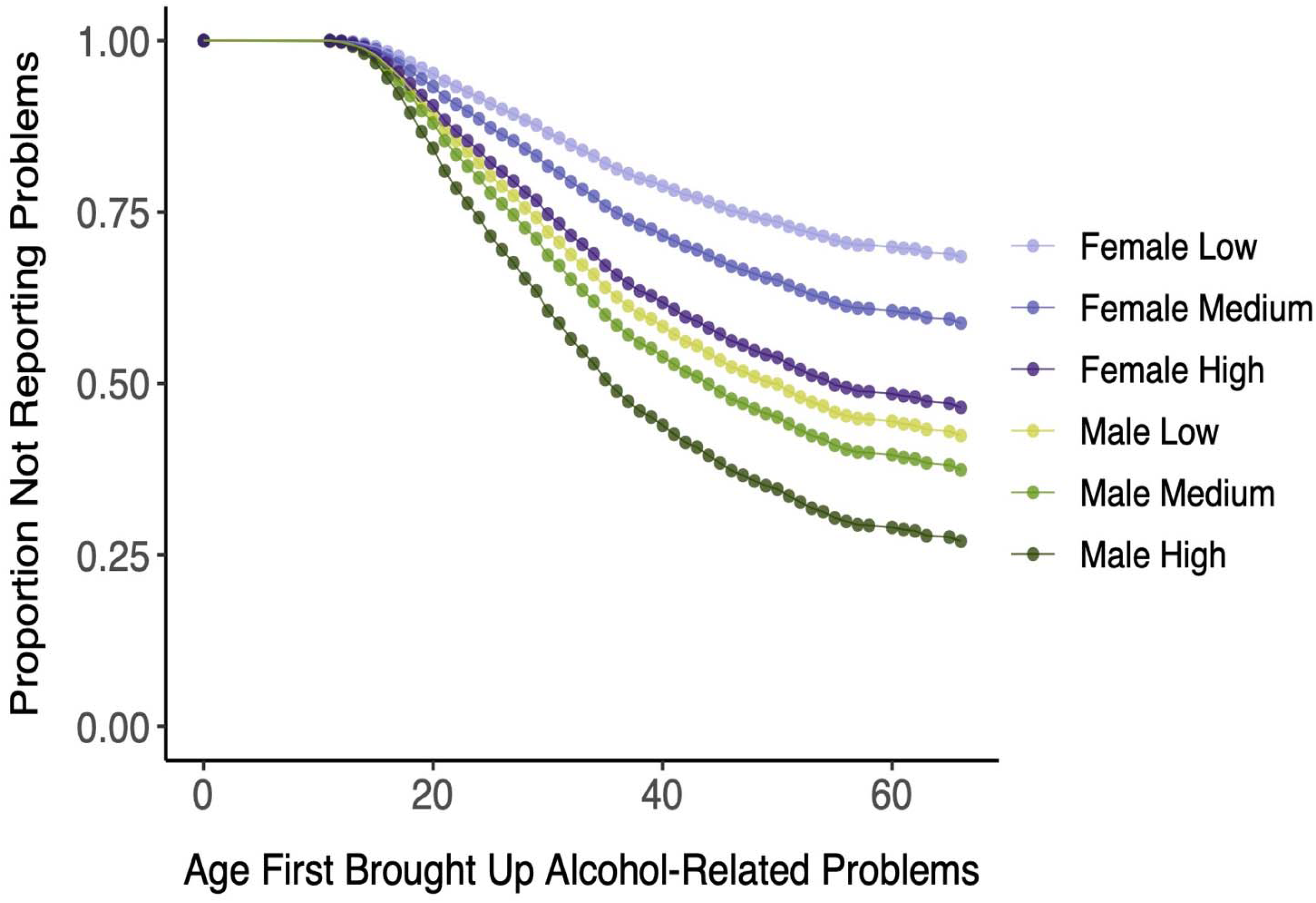
Age First Brought Up Alcohol-Related Problems to a Healthcare Professional Among European-Ancestry Individuals by Sex Alcohol Use Disorder Polygenic Risk Score

Similarly, although the effects of the AUD PRS on the two measures of progression were significant in both sexes, the effects were greater among women (HR=1.27 for progression to reporting problems and 1.14 for progression to AD) than among men (HR=1.09 for both). The progression from regular drinking to first reported alcohol-related problems in EUR differed significantly by sex (*p*_*adj*_=0.0054), with men bringing up alcohol-related problems sooner after beginning regular drinking than women. As can be seen in Figure 4, there is overlap between the survival curves of females in the highest PRS tertile and males in the lowest PRS tertile.

**Figure 4.**
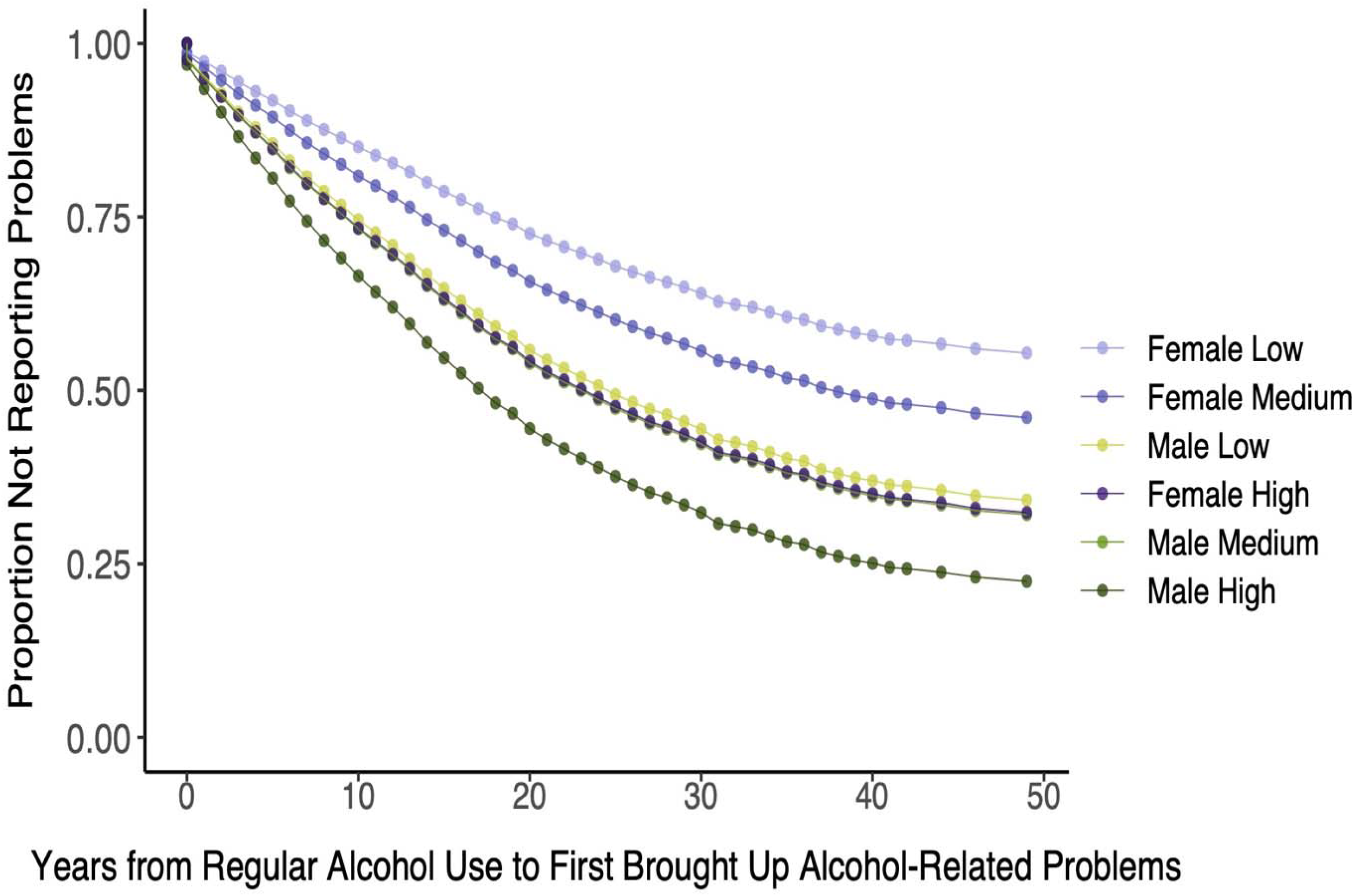
Years from First Regular Alcohol Use to First Brought Up Alcohol-Related Problems to a Healthcare Professional Among European-Ancestry Individuals by Sex Alcohol Use Disorder Polygenic Risk Score

Among AFR, the only significant effect of the alcohol PRS when stratified by sex was on age of AD diagnosis. The effect was significant in both men (HR=1.09) and women (HR=1.11) (Supplemental Table 2).

### Opioid-related Milestones and Progression

Among EUR, the OUD PRS significantly predicted all four milestones (HRs=1.14-1.19) but neither of the progression outcomes. Among AFR, the OUD PRS was not associated with any opioid-related milestones or progression outcomes (Table 5). Figure 1 (middle two panels) shows the difference between population groups in the survival curves for age of first OD diagnosis as a function of OUD PRS strata.

**Table 5.**
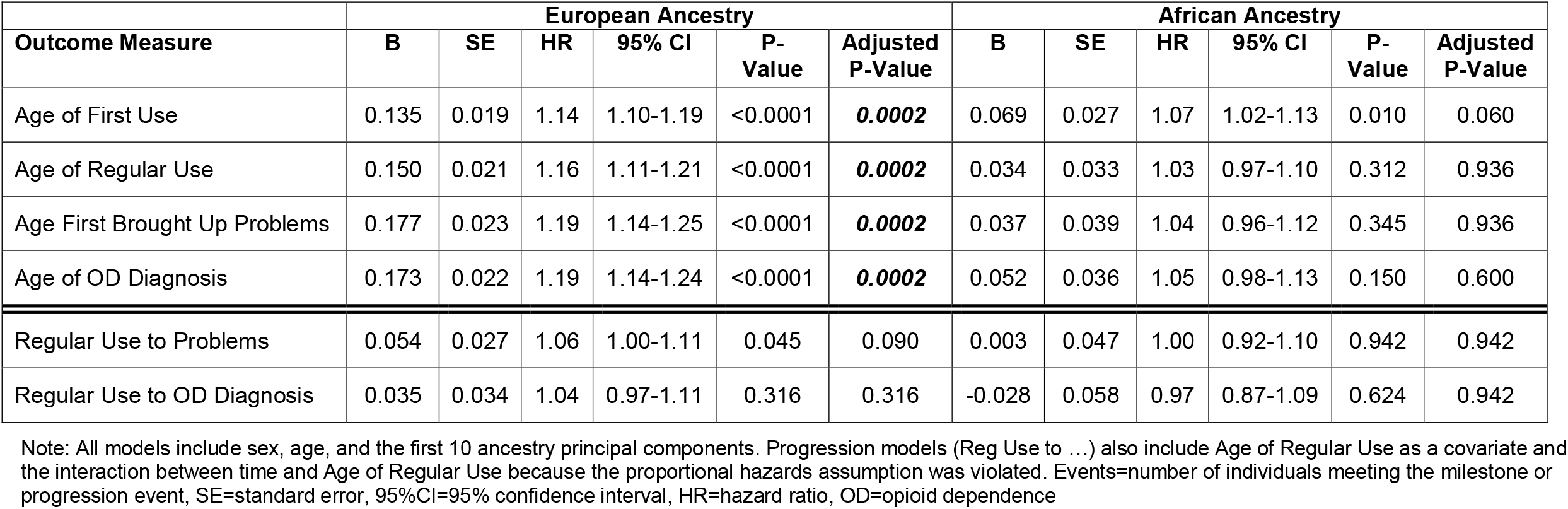
Effect of Opioid Use Disorder Polygenic Risk Score in Cox Proportional Hazard Models of Opioid-Related Time-to-Event and Progression Measures by Population Group

| Outcome Measure | European Ancestry |  |  |  |  |  | African Ancestry |  |  |  |  |  |
| --- | --- | --- | --- | --- | --- | --- | --- | --- | --- | --- | --- | --- |
|  | B | SE | HR | 95% CI | P-Value | Adjusted P-Value | B | SE | HR | 95% CI | P-Value | Adjusted P-Value |
| Age of First Use | 0.135 | 0.019 | 1.14 | 1.10-1.19 | <0.0001 | **0.0002** | 0.069 | 0.027 | 1.07 | 1.02-1.13 | 0.010 | 0.060 |
| Age of Regular Use | 0.150 | 0.021 | 1.16 | 1.11-1.21 | <0.0001 | **0.0002** | 0.034 | 0.033 | 1.03 | 0.97-1.10 | 0.312 | 0.936 |
| Age First Brought Up Problems | 0.177 | 0.023 | 1.19 | 1.14-1.25 | <0.0001 | **0.0002** | 0.037 | 0.039 | 1.04 | 0.96-1.12 | 0.345 | 0.936 |
| Age of OD Diagnosis | 0.173 | 0.022 | 1.19 | 1.14-1.24 | <0.0001 | **0.0002** | 0.052 | 0.036 | 1.05 | 0.98-1.13 | 0.150 | 0.600 |
| Regular Use to Problems | 0.054 | 0.027 | 1.06 | 1.00-1.11 | 0.045 | 0.090 | 0.003 | 0.047 | 1.00 | 0.92-1.10 | 0.942 | 0.942 |
| Regular Use to OD Diagnosis | 0.035 | 0.034 | 1.04 | 0.97-1.11 | 0.316 | 0.316 | -0.028 | 0.058 | 0.97 | 0.87-1.09 | 0.624 | 0.942 |

Note: All models include sex, age, and the first 10 ancestry principal components. Progression models (Reg Use to ...) also include Age of Regular Use as a covariate and the interaction between time and Age of Regular Use because the proportional hazards assumption was violated. Events=number of individuals meeting the milestone or progression event, SE=standard error, 95%CI=95% confidence interval, HR=hazard ratio, OD=opioid dependence

When stratified by sex, among EUR, the OUD PRS was significantly associated in both sexes with all four of the milestones but neither of the progression measures (Supplemental Table 3).

Among AFR, sex-stratified analyses of the effects of the OUD PRS show that the only milestone that was significant was age of onset of opioid use, an effect limited to women (Supplemental Table 4).

### Smoking-related Milestones and Progression

Table 6 shows the results from the Cox models for smoking-related traits. Among EUR, as with OUD, the SMK PRS significantly predicted all four age-of-onset measures (HRs=1.15-1.25), while among AFR it predicted only age of first use (HR=1.05). Survival curves for the age of diagnosis of ND by PRS strata (Figure 1, lower two panels), show the population differences on this key milestone. In neither population group was SMK PRS a significant predictor of the progression from regular smoking to reporting smoking-related problems or to ND.

**Table 6.** Effect of Smoking Polygenic Risk Score in Cox Proportional Hazard Models of Smoking-Related Time-to-Event and Progression Measures by Population Group

| Outcome Measure | European Ancestry |  |  |  |  |  | African Ancestry |  |  |  |  |  |
| --- | --- | --- | --- | --- | --- | --- | --- | --- | --- | --- | --- | --- |
|  | B | SE | HR | 95% CI | P-Value | Adjusted P-Value | B | SE | HR | 95% CI | P-Value | Adjusted P-Value |
| Age of First Use | 0.139 | 0.015 | 1.15 | 1.12-1.18 | <0.0001 | **0.0002** | 0.044 | 0.016 | 1.05 | 1.01-1.08 | 0.006 | **0.036** |
| Age of Regular Use | 0.217 | 0.017 | 1.24 | 1.20-1.29 | <0.0001 | **0.0002** | 0.027 | 0.018 | 1.03 | 0.99-1.07 | 0.125 | 0.446 |
| Age First Brought Up Problems | 0.220 | 0.025 | 1.25 | 1.19-1.31 | <0.0001 | **0.0002** | 0.039 | 0.032 | 1.04 | 0.98-1.11 | 0.223 | 0.669 |
| Age of ND Diagnosis | 0.223 | 0.019 | 1.25 | 1.21-1.30 | <0.0001 | **0.0002** | 0.026 | 0.020 | 1.03 | 0.99-1.07 | 0.188 | 0.564 |
| Regular Use to Problems | 0.039 | 0.026 | 1.04 | 0.99-1.09 | 0.137 | 0.137 | 0.023 | 0.033 | 1.02 | 0.96-1.09 | 0.479 | 0.958 |
| Regular Use to ND Diagnosis | 0.041 | 0.020 | 1.04 | 1.00-1.08 | 0.040 | 0.080 | -0.000 | 0.021 | 1.00 | 0.96-1.40 | 0.980 | 0.980 |

Note: All models include sex, age, and the first 10 ancestry principal components. Progression models (Reg Use to ...) also include Age of Regular Use as a covariate and the interaction between time and Age of Regular Use because the proportional hazards assumption was violated. Events=number of individuals meeting the milestone or progression event, SE=standard error, 95%CI=95% confidence interval, HR=hazard ratio, ND=tobacco dependence

Among EUR, sex-stratified analyses showed that, in both sexes, the SMK PRS is associated with all four milestones and, among females, with the progression from regular smoking to onset of ND (Supplemental Table 5). Similar analyses among AFR (Supplemental Table 6) yielded only one significant effect on smoking-related traits: among males SMK PRS predicted the age of smoking onset.

## Discussion

There is growing interest in the clinical utility of PRS for identifying individuals at high risk for a variety of disorders. In addition to case identification, estimates of genetic risk are increasingly being used to predict disease progression. We used such an approach to demonstrate polygenic effects on the age of onset of substance-related traits and the progression from regular substance use to substance-related problems and dependence. To do this, we used summary statistics from large GWAS of AUD,^20^ OUD,^21^ and SMK^22^ to calculate PRS in a sample of deeply phenotyped individuals either with alcohol and drug use disorders or who were screened controls. We also compared these effects by population group and by sex within population groups.

Our most consistent findings were for alcohol-related traits. Among EUR, an AUD PRS predicted the age of all four alcohol-related milestones and both measures of progression from the onset of regular drinking. The findings replicate a previous observation that an AD PRS predicted the progression from onset of regular drinking to AD diagnosis in a sample of EUR.^18^ We also extended the analysis to AFR, among whom an AUD PRS significantly predicted the age of regular alcohol use, the age of an AD diagnosis, and the progression from regular use to an AD diagnosis.

Stratifying the analyses on the age of onset of regular drinking did not substantially alter the findings. Sex-stratified analyses showed that for some outcomes, the effects of PRS were greater among women than men. These findings could be relevant to the phenomenon of telescoping, in which women who, despite initiating substance use at a later age than men have been reported to have a more rapid progression to developing problems and presenting for treatment than men.^28,29^ However, the findings supporting telescoping are inconsistent (e.g.,^12,30^). Here we found that among EUR the AUD PRS predicted a significantly shorter time from onset of regular alcohol use to bringing up alcohol-related problems among men than women and among AFR the effect of the AUD PRS on the progression from regular drinking to onset of AD was comparable for men and women.

Among EUR, there were also robust effects of OUD and SMK PRS on opioid- and tobacco-related milestones, respectively. However, neither PRS predicted the progression from age of onset of regular use either to bringing up problems related to these substances or dependence diagnoses. Among AFR, the only opioid-related milestone that was significantly associated with OUD PRS was the age of onset of opioid use among women. In this population group, the SMK PRS was associated with an earlier age of smoking initiation, a finding that was significant in men only.

Overall, we found consistently greater associations of PRS with substance-related milestones and symptom progression in EUR than AFR, attributable to the greater predictive value of the EUR summary statistics for all three substances, evidenced by greater genomic inflation factor values among EUR. Further, despite comparable numbers of EUR and AFR individuals with AD and ND in the Yale-Penn (i.e., target) sample, the number of AFR subjects in the target sample that endorsed opioid-related milestones and that met criteria for OD was about one-third the number among EUR. Thus, for opioids, differences in the target sample also likely contributed to the population-group difference in PRS effects. Differences in the sizes of the discovery and target samples by population group underscore the need, particularly in non-EUR populations, for larger GWAS samples and additional deeply phenotyped samples for more granular studies of genetic risk for substance use milestones and progression. This would require targeted recruitment efforts.

Larger discovery samples would increase the predictive power in both population groups, thereby accounting for more variance in the progression to problematic substance use. A more powerful PRS could thus aid in identifying individuals at greatest risk for developing more serious substance-related consequences and permit secondary preventive efforts. One potential test of the clinical utility of PRS in AUD would involve recruiting individuals at the upper end of the polygenic risk continuum for the disorder to participate in clinical trials of interventions aimed at preventing the progression to problematic alcohol use. Existing biobanks could be used to accomplish such an effort.

Among EUR, the effects of the AUD PRS were more consistent and robust than were the OUD or SMK PRS. This could have resulted in part from there being approximately one-third fewer participants in the SMK GWAS than either the AUD or OUD GWAS and approximately one-quarter fewer Yale-Penn participants with an OD diagnosis than either an AD or ND diagnosis. Substance-specific differences have also been shown to exist in symptom progression. National survey data showed that the cumulative probability of progressing to dependence was 67.5% for nicotine users and 22.7% for alcohol users.^31^ A prospective study of adolescents showed that the shortest progression times (i.e., greatest addictive liabilities) were seen with opioids; tobacco and alcohol had the lowest liabilities.^32^ Despite these findings, there are countervailing biological effects. For example, the rate of absorption of nicotine from smoking is much higher than gastrointestinal absorption of alcohol given the extensive surface area of pulmonary alveoli.^33,34^ Thus, pharmacologic or other features specific to individual substances could add to or interact with genetic risk for dependence on them.

Although here we focus principally on genetic risk, environmental factors are also relevant to the age at which substance-related milestones occur.^35^ Among both AFR and EUR, we found the lowest HRs for the age of first alcohol use (1.02 and 1.06, respectively). This is consistent with the notion that the initiation of substance use is strongly influenced by social and environmental factors, whereas the progression from first use to heavy use and from heavy use to problematic use or dependence is influenced more by neurobiological, including genetic, factors.^35^ This is further supported by the finding that the age at first alcohol use is only modestly genetically correlated with AD (r_g_=18–29%),^36^ while for age of onset of regular drinking and AD the genetic correlation is moderate (r_g_=0.54).^37^

This study has limitations. We conducted analyses in only two population groups—EUR and AFR—the smaller numbers of Hispanic, East Asian, and South Asian individuals in both the discovery and target samples do not provide adequate statistical power to support these analyses. We also used DSM-IV SUD diagnoses in the target sample to ensure consistency across the substances, as we did not have all criteria required for a DSM-5 tobacco use disorder diagnosis. Third, we used a trajectory phenotype in the discovery GWAS for smoking, as it was the largest available GWAS for smoking in AFR. Nonetheless, the phenotype differs from the ICD-9/10 codes used in the AUD and OUD GWAS. Whereas the trajectories are a probabilistic categorization rather than a binary diagnosis, the trajectory-based groups are potentially more heterogeneous than AUD or OUD cases and controls. This variability could have diminished the association of the SMK PRS with smoking-related milestones or latency outcomes. Finally, the proportions of SUD diagnoses overall and by population reflect the recruitment strategy of the original studies, and therefore no conclusions may be drawn about these proportions *per se*.

Despite these limitations, the availability of large discovery GWAS and the deeply phenotyped target sample made it possible to generate sex- and population-specific profiles of the association of polygenic risk for SUDs with both milestones and symptom progression over time. These findings provide an initial characterization of the genetic risk of these more granular features and a foundation for efforts to identify higher-risk individuals who can be targeted with interventions aimed at preventing symptom progression.

## Data Availability

All data produced in the present study are available upon reasonable request to the authors

## Acknowledgments

Supported by the Veterans Integrated Service Network 4 Mental Illness Research, Education and Clinical Center and NIH grants P30 DA046345, R01 AA026364, and K01 AA028292 (to RLK).

