## Supplemental text, tables, and figures for "Polygenic Risk for Substance-Related Traits Predicts Substance Use Onset and Progression: Sex and Population Group Differences"

**Supplemental Methods**

The discovery sample for AUD (Kember et al. 2022a) comprised 296,989 EAs and 80,764 AAs from the Million Veteran Program (MVP), the largest AUD sample currently available. The mean age among EAs was 62 (standard deviation (SD) = 13) and among AAs, it was 56 (SD = 12). Most individuals in both population groups were male (EA: 92.7% and AA: 86.8%). Cases were individuals who received at least one inpatient or two outpatient ICD-9/10 diagnostic codes for AUD. This consisted of 63,375 EA cases (21.3% of the total) and 28,541 AA cases (35.2% of the total).

The discovery sample for OUD (Kember et al. 2022b) included 302,585 EAs and 88,498 AAs from MVP, the GWAS sample that has yielded the greatest number of genome-wide loci to date. The mean age among EAs was 63 (SD = 13) and among AAs, it was 58 (SD = 12). Most individuals in both population groups were male (EA: 92% and AA: 86%). Individuals with at least 1 ICD-9/10 code for opioid abuse or dependence were considered cases. Exposed controls were individuals with one or more outpatient opioid prescription fill. Among EAs, there were 19,978 (7%) cases and 282,607 (93%) controls and among AAs, 8,968 (10%) cases and 79,530 (90%) controls.

The SMK discovery sample (Xu et al. 2020) included 286,118 individuals (209,915 EAs and 54,867 AAs) from MVP. This GWAS differentiated cases from controls using smoking trajectories and is the largest for which matched ancestry summary statistics for AAs are currently available. The mean age among EAs was 64 (SD = 13) and among AAs, it was 58 (SD = 12). Most members of both population groups were male (EA: 93% and AA: 87%). Cases were those with smoking trajectories categorized as “mostly current smokers,” whereas controls’ trajectories were categorized as “mostly never” (see Xu et al, 2020). When calculating the SMK PRS, we excluded individuals in the “mixed” smoking trajectory group. Among EAs, there were 40,456 (19%) cases and 59,056 (28%) controls and among AAs, 13,511 (25%) cases and 17,751 (32%) controls.

**Supplemental Tables**

Supplemental Table 1: Effect of Stratifying on Age of Regular Alcohol Use on the Association of Alcohol Use Disorder Polygenic Risk Score on Alcohol-Related Progression Measures by Population Group

|  | **African Ancestry** | | | | | **European Ancestry** | | | | |
| --- | --- | --- | --- | --- | --- | --- | --- | --- | --- | --- |
| **Outcome Measure** | **B** | **SE** | **HR** | **95% CI** | **P-Value** | **B** | **SE** | **HR** | **95% CI** | **P-Value** |
| **Regular Use to Problems**      Early Onset (≤18 yr)      PRS | -0.032  0.019 | 0.015  0.033 | 0.97  1.02 | 0.94-0.99  0.96-1.09 | ***0.026***  0.556 | -0.080  0.124 | 0.013  0.026 | 0.92  1.13 | 0.90-0.95  1.08-1.19 | ***<0.0001***  ***<0.0001*** |
| Late Onset (>18 yr)       PRS | 0.036  0.087 | 0.012  0.055 | 1.04  1.09 | 1.01-1.06  0.98-1.22 | ***0.004***  0.113 | 0.031  0.168 | 0.016  0.053 | 1.03  1.18 | 1.00-1.06  1.07-1.31 | 0.051  ***0.002*** |
| **Regular Use to AD Diagnosis**      Early Onset (≤18 yr)      PRS | -0.021  0.053 | 0.012  0.028 | 0.98  1.05 | 0.96-1.00  1.00-1.11 | 0.089  0.061 | -0.050  0.090 | 0.011  0.023 | 0.95  1.09 | 0.93-0.97  1.05-1.14 | ***<0.0001***  ***<0.0001*** |
| Late Onset (>18 yr)       PRS | 0.002  0.107 | 0.010  0.044 | 1.00  1.11 | 0.98-1.02  1.02-1.21 | 0.834  ***0.014*** | 0.029  0.110 | 0.012  0.044 | 1.03  1.12 | 1.01-1.05  1.02-1.22 | ***0.016***  ***0.013*** |

Supplemental Table 2: Effect of Alcohol Use Disorder Polygenic Risk Score in Cox Proportional Hazard Models of Alcohol-Related Time-to-Event and Progression Measures by Sex among Individuals of African Ancestry

| **Outcome** | **Male** | | | | | | |  | **Female** | | | | | |
| --- | --- | --- | --- | --- | --- | --- | --- | --- | --- | --- | --- | --- | --- | --- |
|  | **Events** | **B** | **SE** | **HR** | **95% CI** | **P-**  **Value** | **Adjusted**  **P-Value** | **Events** | **B** | **SE** | **HR** | **95% CI** | **P-**  **Value** | **Adjusted P-Value** |
| Age of First Use | 2,582 | 0.019 | 0.024 | 1.02 | 0.97-1.07 | 0.410 | 0.452 | 2,097 | 0.021 | 0.025 | 1.02 | 0.97-1.07 | 0.400 | 0.400 |
| Age of Regular Use | 2,390 | 0.039 | 0.024 | 1.04 | 0.99-1.09 | 0.105 | 0.320 | 1,644 | 0.051 | 0.029 | 1.05 | 1.00-1.11 | 0.075 | 0.300 |
| Age First Brought Up Problems | 1219 | 0.048 | 0.034 | 1.05 | 0.98-1.12 | 0.160 | 0.452 | 659 | 0.111 | 0.047 | 1.12 | 1.02-1.22 | *0.017* | 0.085 |
| Age of AD Diagnosis | 1,693 | 0.083 | 0.029 | 1.09 | 1.03-1.15 | *0.004* | ***0.024*** | 938 | 0.104 | 0.039 | 1.11 | 1.03-1.20 | *0.007* | ***0.042*** |
| Regular Use to Problems | 1,144 | 0.026 | 0.035 | 1.03 | 0.96-1.10 | 0.452 | 0.452 | 603 | 0.061 | 0.050 | 1.06 | 0.96-1.17 | 0.220 | 0.400 |
| Regular Use to AD Diagnosis | 1,475 | 0.060 | 0.031 | 1.06 | 1.00-1.13 | *0.050* | 0.250 | 778 | 0.072 | 0.044 | 1.07 | 0.99-1.17 | 0.102 | 0.306 |

Supplemental Table 3: Cox Regression Models for Opioid Use Disorder Polygenic Risk Score Prediction of Opioid-related Time-to-Event and Progression Measures by Sex among Individuals of European Ancestry

| **Outcome** | **Male** | | | | | | **Female** | | | | | |
| --- | --- | --- | --- | --- | --- | --- | --- | --- | --- | --- | --- | --- |
|  | **B** | **SE** | **HR** | **95% CI** | **P-Value** | **Adjusted P-Value** | **B** | **SE** | **HR** | **95% CI** | **P-Value** | **Adjusted P-Value** |
| Age of First Use | .145 | .023 | 1.16 | 1.10-1.21 | <0.0001 | ***0.0002*** | .125 | .033 | 1.13 | 1.06-1.21 | 0.0001 | ***0.0003*** |
| Age of Regular Use | .144 | .026 | 1.16 | 1.10-1.22 | <0.0001 | ***0.0002*** | .167 | .037 | 1.18 | 1.10-1.27 | <0.0001 | ***0.0002*** |
| Age First Brought Up Problems | .175 | .028 | 1.19 | 1.13-1.26 | <0.0001 | ***0.0002*** | .188 | .039 | 1.21 | 1.12-1.30 | <0.0001 | ***0.0002*** |
| Age of OD Diagnosis | .170 | .027 | 1.19 | 1.12 -1.25 | <0.0001 | ***0.0002*** | .181 | .037 | 1.20 | 1.11-1.29 | <0.0001 | ***0.0002*** |
| Regular Use to Problems | .068 | .033 | 1.07 | 1.00-1.14 | 0.040 | 0.080 | .031 | .046 | 1.03 | 0.94-1.13 | .501 | .501 |
| Regular Use to OD Diagnosis | .059 | .042 | 1.06 | 0.98-1.15 | 0.156 | 0.156 | -.069 | .064 | 0.93 | 0.82-1.06 | .375 | .501 |

Supplemental Table 4: Cox Regression Models for Opioid Use Disorder Polygenic Risk Score Prediction of Opioid-related Time-to-Event and Progression Measures by Sex among Individuals of African Ancestry

| **Outcome** | **Male** | | | | | | **Female** | | | | | |
| --- | --- | --- | --- | --- | --- | --- | --- | --- | --- | --- | --- | --- |
|  | **B** | **SE** | **HR** | **95% CI** | **P-Value** | **Adjusted P-Value** | **B** | **SE** | **HR** | **95% CI** | **P-Value** | **Adjusted P-Value** |
| Age of First Use | .041 | .033 | 1.04 | 0.98-1.10 | .211 | .960 | .135 | .047 | 1.15 | 1.04-1.26 | .004 | ***.024*** |
| Age of Regular Use | -.004 | .041 | 1.00 | 0.92-1.08 | .925 | .960 | .116 | .057 | 1.12 | 1.00-1.26 | .043 | .129 |
| Age First Brought Up Problems | -.005 | .049 | 1.00 | 0.90-1.10 | .927 | .960 | .126 | .064 | 1.13 | 1.00-1.29 | .051 | .153 |
| Age of OD Diagnosis | .025 | .045 | 1.03 | 0.94-1.12 | .572 | .960 | .120 | .061 | 1.13 | 1.00-1.27 | .048 | .144 |
| Regular Use to Problems | .003 | .058 | 1.00 | 0.89-1.12 | .960 | .960 | -.031 | .086 | 0.97 | 0.82-1.15 | .718 | .718 |
| Regular Use to OD Diagnosis | .014 | .068 | 1.01 | 0.89-1.16 | .841 | .960 | -.183 | .112 | 0.83 | 0.67-1.04 | .104 | .208 |

  Supplemental Table 5: Cox Regression Models for Smoking Polygenic Risk Score Prediction of Smoking-related Time-to-Event and Progression Measures by Sex among Individuals of European Ancestry

| **Outcome** | **Male** | | | | | | **Female** | | | | | |
| --- | --- | --- | --- | --- | --- | --- | --- | --- | --- | --- | --- | --- |
|  | **B** | **SE** | **HR** | **95% CI** | **P-Value** | **Adjusted P-Value** | **B** | **SE** | **HR** | **95% CI** | **P-Value** | **Adjusted P-Value** |
| Age of First Use | 0.130 | 0.020 | 1.14 | 1.01-1.18 | <0.0001 | ***0.0002*** | 0.152 | 0.023 | 1.16 | 1.11-1.22 | <0.0001 | ***0.0002*** |
| Age of Regular Use | 0.191 | 0.022 | 1.21 | 1.06-1.26 | <0.0001 | ***0.0002*** | 0.258 | 0.028 | 1.29 | 1.22-1.37 | <0.0001 | ***0.0002*** |
| Age First Brought Up Problems | 0.182 | 0.032 | 1.20 | 1.13-1.28 | <0.0001 | ***0.0002*** | 0.283 | 0.041 | 1.33 | 1.22-1.44 | <0.0001 | ***0.0002*** |
| Age of OD Diagnosis | 0.176 | 0.024 | 1.19 | 1.14-1.25 | <0.0001 | ***0.0002*** | 0.310 | 0.031 | 1.36 | 1.28-1.45 | <0.0001 | ***0.0002*** |
| Regular Use to Problems | 0.028 | 0.033 | 1.03 | 0.96-1.10 | 0.402 | 0.787 | 0.068 | 0.042 | 1.07 | 0.99-1.16 | 0.106 | 0.106 |
| Regular Use to OD Diagnosis | 0.007 | 0.025 | 1.01 | 0.96-1.06 | 0.787 | 0.787 | 0.100 | 0.033 | 1.11 | 1.04-1.18 | 0.002 | ***0.004*** |

Supplemental Table 6: Cox Regression Models for Smoking Polygenic Risk Score Prediction of Smoking-related Time-to-Event and Progression Measures by Sex among Individuals of African Ancestry

| **Outcome** | **Male** | | | | | | **Female** | | | | | |
| --- | --- | --- | --- | --- | --- | --- | --- | --- | --- | --- | --- | --- |
|  | **B** | **SE** | **HR** | **95% CI** | **P-Value** | **Adjusted P-Value** | **B** | **SE** | **HR** | **95% CI** | **P-Value** | **Adjusted P-Value** |
| Age of First Use | 0.058 | 0.021 | 1.06 | 1.02-1.11 | 0.007 | ***0.042*** | 0.026 | 0.024 | 1.03 | 0.98-1.08 | 0.281 | 0.768 |
| Age of Regular Use | 0.046 | 0.023 | 1.05 | 1.00-1.10 | 0.051 | 0.204 | 0.005 | 0.028 | 1.01 | 0.95-1.06 | 0.848 | 0.953 |
| Age First Brought Up Problems | 0.007 | 0.043 | 1.01 | 0.93-1.10 | 0.867 | 0.867 | 0.092 | 0.049 | 1.10 | 1.00-1.21 | 0.062 | 0.372 |
| Age of OD Diagnosis | 0.048 | 0.026 | 1.05 | 1.00-1.10 | 0.068 | 0.272 | -0.002 | 0.031 | 1.00 | 0.94-1.06 | 0.953 | 0.953 |
| Regular Use to Problems | 0.008 | 0.045 | 1.01 | 0.92-1.10 | 0.851 | 0.867 | 0.043 | 0.050 | 1.05 | 0.95-1.15 | 0.385 | 0.768 |
| Regular Use to OD Diagnosis | 0.015 | 0.027 | 1.02 | 0.96-1.07 | 0.576 | 0.867 | -0.021 | 0.032 | 0.98 | 0.92-1.04 | 0.512 | 0.768 |
